# Nurses’ Perceptions regarding the antiretroviral therapy services at selected health facilities in Lesotho

**DOI:** 10.1101/2021.10.21.21265325

**Authors:** Isabel Nyangu, Zerish Zethu Nkosi

**Affiliations:** Department of Nursing, National University of Lesotho, P. O. Roma 180, Lesotho; College of Human Sciences, College of Human Sciences, Muckleuneck Ridge, Pretoria 0002, South Africa

## Abstract

**Background:** In Lesotho, ART services are provided in primary health care (PHC) facilities which are mostly run by nurses.

**Purpose:** The study aimed to describe perceptions of nurses regarding the antiretroviral therapy services at PHC facilities across six selected districts in Lesotho.

**Methods:** A cross sectional survey was conducted in which convenience sampling was used to select 214 nurses working at PHC facilities across six districts. They completed a structured self-report questionnaire that collected information on their perceptions using a Likert scale with six options (strongly agree, agree, neutral, disagree, strongly disagree, and no answer). The response rate was 92% (*n*=197) and data were analyzed using SPSS (23).

**Results:** Nurses’ perceptions were positive and the same on whether ARVS and other medications were available; they were qualified and competent to provide ART services; they had adequate resources and equipment to provide ART services; they were satisfied with their jobs and the services they provided; and there was monitoring and evaluating system for the ART services. Nurses’ perceptions significantly differed and were mostly negative on whether they were adequately staffed to provide ART services; their organizational structures allowed for the provision of adequate and efficient services; ART services were accessible; and there was adequate and accurate documentation of the ART services they provided.

**Conclusions:** As nurses are the main health care providers in Lesotho, their perceptions are important in improving service delivery. It is important to standardize ART services across the districts to ensure equitable accessibility in line with service demand.

## Introduction

Lesotho has one of the highest human immunodeficiency virus (HIV) prevalence (23.8%) among adult men and women aged15-49 years which is a major developmental challenge for the country (UNAIDS, 2017). It is estimated that 306 000 adults aged above 15 years and 13 000 children aged 0 to 14 years are living with HIV in Lesotho (LePHIA, 2019). The national HIV incidence among adults (15-49 years) is 1.55%, with women having a higher incidence (1.81%) compared to men at 1.33% (National HIV & AIDS Strategic Plan (NHASP) 2018/19-2022/23). Lesotho initiated antiretroviral therapy in 2001 and adopted the test and treat strategy in 2016 (LePHIA, 2019). Additionally, the country has achieved 77-90-88 targets amongst people living with HIV/AIDS, with 88% of those on antiretroviral therapy (ART) reportedly virally suppressed (LePHIA, 2019). Unfortunately, 11.4% of adults on ART have developed transmitted resistance to non-nucleoside reverse transcriptase inhibitors (NNRTIs), a pattern suggestive of treatment failure due to pre-existing drug resistance (NHSAP 2018/19-2022/23). Apparently, one doctor and three nurses serve a population of 1000 people living with HIV or Tuberculosis (TB) and the majority of the health facilities are staffed as per minimum staffing requirements (LePHIA, 2019). Majority of health care facilities in Lesotho are managed by nurses who are qualified as either midwives or clinicians (Nyangu & Nkosi, 2019a). Lesotho is one of the few countries that permitted initiation of ART by nurses owing to the scarcity of Medical Doctors, a move that has seen a great increase of the coverage of ART (Nyangu, 2016). Such an initiative has been seen to be operative and feasible as majority of patients receive health care in the facilities located variously across the country.

However, the geographic topography and limited human resource contribute to inadequate service uptake and delivery (NHASP 2018/19-2022/23). Staff shortages and tedious documentation have been reported to affect service delivery, whilst ART services were offered on varying days across districts and patient waiting times were variable depending on the required services (Nyangu and Nkosi, 2019a). It therefore remains imperative to support nurses and reduce patients waiting times so that the ART program is a success.

### Problem Statement

Lesotho remains highly burdened with HIV/AIDS, with almost 24% of the population infected by the disease. Whilst ART is generally available to the many that need it, there are challenges in its accessibility across the country owing to the differences in the topography and population distribution. Nurses remain the key health care providers in resource limited countries such as Lesotho. Whilst several studies attest to the roles of nurses in the provision of ART services, information on their perceptions regarding the ART services remains limited, especially in Lesotho. This study therefore envisaged addition to the existing body of knowledge on the perceptions of nurses regarding ART services at primary health care (PHC) facilities across selected districts in Lesotho.

## Methods

### Study Design

A cross-sectional survey using structured self-report questionnaire was used to collect data from 214 nurses who were conveniently sampled at PHC facilities across six out of 10 districts in Lesotho. The questionnaire collected information of the perceptions of nurses using a Likert scale with six options (strongly agree, agree, neutral, disagree, strongly disagree, and no answer). A pilot survey was conducted to ascertain relevance of items on the questionnaire (content validity). Cronbach’s Alpha coefficient was used to measure internal consistency and it was 0.785. SPSS (version 23) was used to analyze data. The compliance rate was 92% with 197 respondents out of the expected 214 responding to the questionnaire.

### Data collection

As they convened for their monthly planning meetings, researchers introduced the study to nurses. Upon agreeing to participate, the nurses completed an informed consent form and structured questionnaire. Each nurse completed the questionnaires privately and could ask clarity seeking questions to the researcher.

### Ethical Considerations

Benchmarks for ethical research by Emmanuel (Emanuel *et al*., 2004) were used in this study. Permission to undertake the study was granted from relevant ethics bodies (REC-012714; ID136-2014). Respondents were asked to give written consent to acknowledge their voluntary participation in the study. Only codes were used to identify respondents and those who declined to participate or withdrew from the study were treated in a non-judgmental way. They were allowed to ask questions, refuse to give information, and there were no implicit or explicit threats (coercion) of penalty from failing to participate or excessive rewards from agreeing to participate.

## RESULTS

### Demographic Characteristics of Respondents

Seven percent (*n*=14) of the respondents were males and 92% (*n*=181) were females. The mean age of the respondents was 36 years (*CI*=34.8-38.3). Twenty-nine percent (*n*=36) of the respondents’ were aged 20-29 years, 43% (*n*=54) were aged 30-39 years, 14% (*n*=18) were aged 40-49 years, 11% (*n*=14) were aged 50-59 years, and 3% (*n*=4) were aged 60-69 years. Majority of the PHC facilities were staffed by nurse midwives (72%; *n*=142) and nurse clinicians (19%; *n*=37).

### Perceptions of nurses regarding the ART services

#### Antiretroviral therapy medication

Most of the nurses had positive perceptions that ARVs were always available at their facilities (66% (*n*=130) strongly agreed, and 27% (*n*=54) agreed). Only a few had negative perceptions (1% (*n*=2) disagreed, and 2% (*n*=4) strongly disagreed), whereas 3% (*n*=5) were neutral. The responses were analyzed using Kruskal-Wallis ANOVA (KWA) and the result was not significant (*p*=0.334; α=0.05). It was concluded that perceptions on the availability of ARVs were the same across the selected districts in Lesotho.

#### Medication to manage other health conditions

Majority of the nurses had positive perceptions that medicines to manage other health conditions were always available (24% (*n*=47) strongly agreed and 51% (*n*=101) agreed). Minority of the respondents had negative perceptions (12% (*n*=24) disagreed, 1% (*n*=2) strongly disagreed), whilst 12% (*n*=23) were neutral. The responses were analyzed using KWA and the result was not significant (*p*=0.447; α=0.05). The conclusion was that perceptions on the availability of medicines to manage other health conditions were the same across the selected districts in Lesotho.

#### Staffing

Some of the nurses had positive perceptions that their facilities had adequate staff to provide ART services (16% (*n*=32) strongly agreed and 23% (*n*=45) agreed). Other nurses had negative perceptions (28% (*n*=55) disagreed and 19.3% (*n*=38) strongly disagreed), whilst 13.2% (*n*=26) were neutral. The responses were analyzed using KWA and the result was significant (*p*=0.000; α=0.05). It was concluded that perceptions on the adequacy of staff to provide ART services were significantly different across the districts.

#### Competency of staff

Majority of the nurses had positive perceptions that staff providing ART services were qualified and competent (58% (*n*=114) strongly agreed, 31% (*n*=62) agreed) to provide ART services. A few nurses had negative perceptions (9% (*n*=17) disagreed), and 1.5% (*n*=3) were neutral. The responses were analyzed using KWA and the result was not significant (*p*=0.305 α=0.05). The conclusion was that perceptions on qualification and competency of staff to provide ART services were the same at facilities across the selected districts.

#### Resources and equipment

Most of the nurses had positive perceptions that their facilities had adequate resources and equipment for the provision of ART services (28% (*n*=58) strongly agreed and 42% (*n*=82) agreed). Only a few nurses had negative perceptions (9% (*n*=17) disagreed and 5.5% (*n*=11) strongly disagreed), whilst 15% (*n*=30) were neutral. The responses were analyzed using KWA and the result was not significant (*p*=0.153: α=0.05). It was concluded that perceptions on the adequacy of resources and equipment to provide ART services at facilities were the same across the selected districts.

#### Organizational Structure

Majority of the nurses had positive perceptions that their organizational structure allowed for the provision of adequate and efficient ART services (35% (n=68) strongly agreed and 30% (n=60) agreed). A minority of nurses had negative perceptions (10% (n=20) disagreed, 5.5% (n=11) strongly disagreed), whilst 19% (n=37) were neutral. The responses were analyzed using KWA and the result was significant (p=0.047: α=0.05). The conclusion was that perceptions whether the organizational structures allowed for the provision of adequate and efficient services significantly differed across the selected districts.

#### Antiretroviral therapy services

Most nurses had positive perceptions that services were routinely, accurately and efficiently provided to allow accessibility (48% (*n*=94) strongly agreed and 42% (*n*=84) agreed). A few nurses had negative perceptions (6% (*n*=11) disagreed), whilst 4% (*n*=8) were neutral. The responses were analyzed using KWA and the result was significant (*p*=0.003: α=0.05). It was concluded that perceptions on the accessibility of services at facilities significantly differed across the six districts.

#### Documentary evidence

Majority of the nurses had positive perceptions that documentary evidence at their facilities was adequate and accurate (31% (*n*=60) strongly agreed and 45% (*n*=89) agreed). A few nurses had negative perceptions (2.5% (*n*=5) disagreed and 2.5% (*n*=5) strongly disagreed), whilst 19% (*n*=38) were neutral. KWA was used to analyze the responses and the result was significant (*p*=0.020; α=0.05). The conclusion was that perceptions on the adequacy and accuracy of documentary evidence significantly differed across the selected districts.

#### Job satisfaction

Most of the nurses had positive perceptions that they were satisfied with their jobs and ART services offered at health care facilities (34% (*n*=66) strongly agreed and 44% (*n*=86) agreed). A minority of nurses had negative perceptions (12% (*n*=24) disagreed and 3% (*n*=7) strongly disagreed), whilst 7% (*n*=13) were neutral. KWA was used to analyze the responses and the result was not significant (*p*=0.164; α=0.05). It was concluded that nurses’ perceptions on satisfaction with their jobs and ART services offered at health facilities were the same across the selected districts.

#### Monitoring and Evaluation System

Majority of the nurses had positive perceptions that there was a monitoring and evaluating system for the ART program (25% (*n*=48) strongly agreed and 36% (*n*=71) agreed). A few nurses had negative perceptions (6% (*n*=12) disagreed, and 2% (*n*=4) strongly disagreed), whilst 8% (*n*=16) were neutral. KWA was used to analyze the responses and the result was not significant (*p*=0.929: α=0.05). The conclusion was that perceptions on the availability of an efficient monitoring and evaluation system of the ART program were the same across the selected districts. Figure one below further illustrates the results.

**Figure 1:**
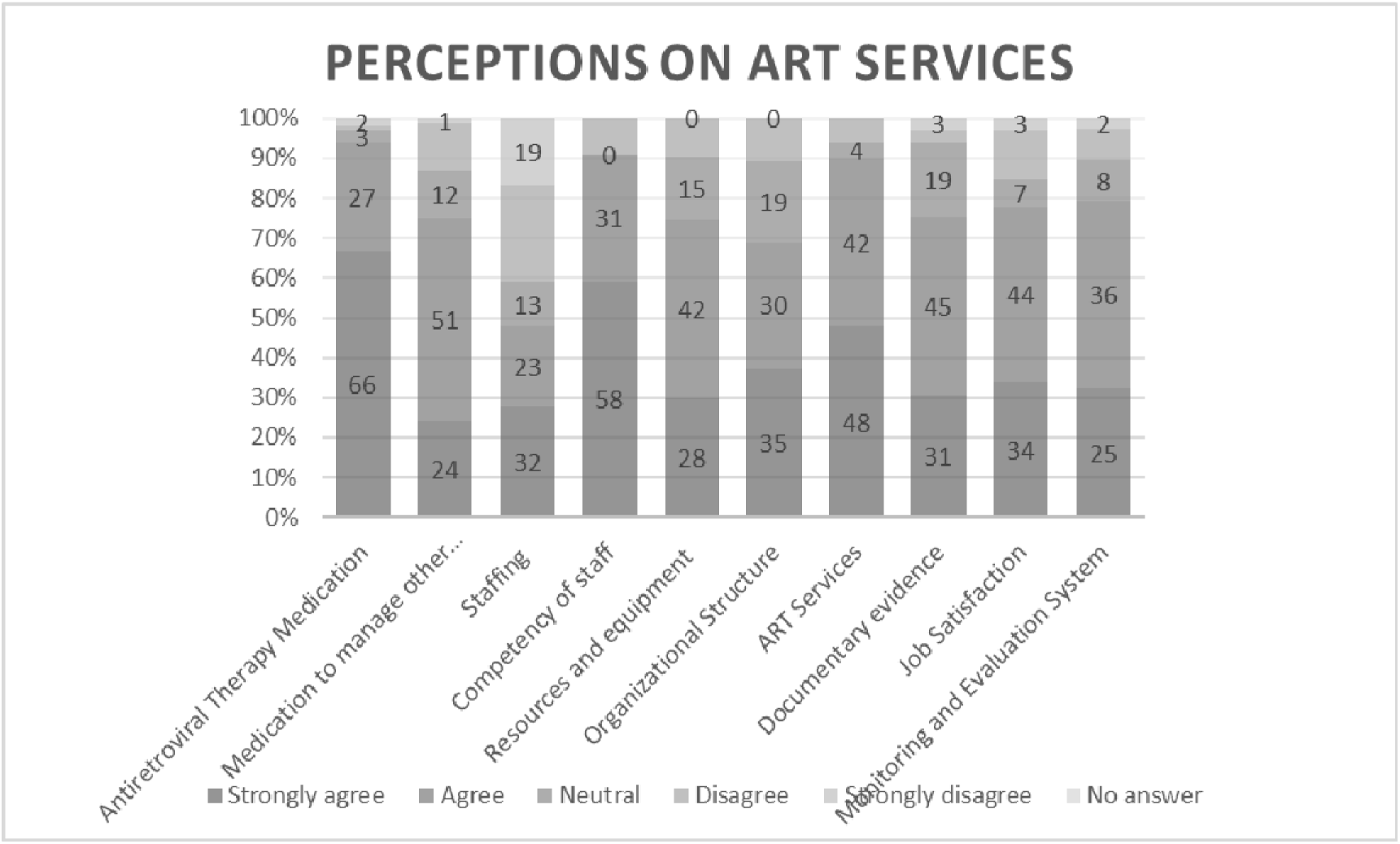
Perceptions on ART Services.

## Discussion

The nurses perceived ARVS and other medications to be generally available at facilities and these perceptions were the same across the selected districts. The availability of medications is important to avoid treatment interruption and improve retention in care. This is in line with Tuhadeleni et.al (Tuhadeleni and Lukolo, 2017) who perceived adherence to be important for the success of ART. Furthermore, retaining patients in care shows progress towards achieving 90-90-90 targets (2017 UNAIDS, 2017) and also results in viral load suppression (Brown *et al*., 2016).

Nurses’ opinions that they were qualified and competent to provide ART services at their facilities were positive and the same across the selected districts. This is important as health care workers are able to assist and monitor patient treatment. This finding is in line with the study by Tuhadeleni et.al (Tuhadeleni and Lukolo, 2017) in which health care providers had positive perceptions about their roles in ART adherence through pill counts, counseling, and questioning. However, the need for supervision remains a challenge in resource limited settings including Lesotho. Mavhandu-Mudzusi et al (Mavhandu-Mudzusi, Sandy and Hettema, 2017) concluded that nurses needed continued support to ensure effective implementation of the ART program and this can increase best practice in healthcare (Rouleau *et al*., 2019).

Perceptions of the nurses that resources and equipment were adequate for the provision of ART services were positive and the same across the districts. Having resources and equipment available improves service delivery which culminates to satisfaction by both patients and nurses. Nurses’ opinions on their job satisfaction were also positive and the same across the selected districts. Job satisfaction culminates to good service delivery and that client service provision remains accurate and adequate, respectively.

Perceptions of nurses on the availability of a monitoring and evaluation system for the ART services were positive and the same across the selected districts in this study. Studies relating monitoring and evaluation of the ART services still remain limited with the Lesotho Demographic and Health survey being the main source of health data and it is conducted every five years. The monitoring and evaluation of health programs is important to identify gaps which will inform improvement strategies.

Perceptions on the adequacy of staffing differed across the districts. Whilst some nurses perceived that they were adequately staffed at their facilities, some opined that they were not. The differences could be the result of different patient populations and staffing patterns across the districts. The shortage of staff continues to be one of the major barriers to effective ART services. Uys and Kopper (Uys and Klopper, 2013) reported that one nurse specialist, five registered nurse midwives, and four enrolled nurses were needed whilst in a study by Nyangu and Nkosi (Nyangu and Nkosi, 2019b) four to five nurses were deemed necessary. Staffing patterns therefore need to be done according to the patient population across the districts.

This study found different perceptions regarding whether the organizational structures allowed for the provision of adequate and efficient ART services across the districts. Some of the nurses perceived that ART services were offered adequately and efficiently as the structure of their organizations allowed them to do so, whilst others did not feel the same way. Organizational structures that support provision of adequate and efficient health care services are crucial to improve retention in care and the lives of those on ART as they are adequately monitored.

Perceptions on the accessibility of ART services differed across the selected districts. Whilst some of the nurses were of the opinion that ART services were accessible, others felt that they were not. These differences could be attributed to the topography of Lesotho, different patient populations, and different staffing patterns across the selected districts. Accessible ART services will result in improved lives of people living with HIV and reduce the HIV disease burden. Similarly, perceptions the adequacy and accuracy of documentary evidence of the ART services were different across the selected districts. Whilst some nurses were of the opinion that they had adequate and accurate documentation on ART services, others did not feel the same way. Documentation of health care service delivery provides an audit trail that can be tracked for identification of gaps and hence inform strategies for improvement. Without adequate documentation of services, it becomes difficult to track the progress of patients on treatment adherence.

## Conclusions

We conclude that nurses had both positive and negative perceptions regarding the ART services. Their perceptions were positive and the same that ARVS and other medications were available; they were qualified and competent to provide ART services; they had adequate resources and equipment to provide ART services; they were satisfied with their jobs and the services they provided; and there was monitoring and evaluating system for the ART services. Nurses’ perceptions differed on whether they were adequately staffed to provide ART services; their organizational structures allowed for the provision of adequate and efficient services; ART services were accessible; and there was adequate and accurate documentation of the ART services they provided. We recommend that ART services be standardized across the districts to ensure equitable accessibility that is in line with service demand.

## Limitations

This study only described perceptions of nurses regarding the ART services at selected health facilities and did not seek to describe outcome impacts of ART services.

## Data Availability

All data produced in the present study are available upon reasonable request to the authors

